# Concurrent prediction of in-hospital mortality and length of stay using single-task, multi-class, and multi-task machine learning

**DOI:** 10.1101/2025.03.21.25324386

**Authors:** Jia Wei, Jo Gay, Andrew J Brent, David A Clifton, A. Sarah Walker, David W. Eyre

**Affiliations:** Nuffield Department of Medicine, University of Oxford, Oxford, UK; Oxford University Hospitals NHS Foundation Trust, Oxford, UK; Department of Engineering Science, University of Oxford, Oxford, UK; The National Institute for Health Research Health Protection Research Unit in Healthcare Associated Infections and Antimicrobial Resistance at the University of Oxford, Oxford, UK; National Institute for Health Research Oxford Biomedical Research Centre, Oxford, UK; Big Data Institute, Nuffield Department of Population Health, University of Oxford, Oxford, UK

## Abstract

**Background:** Accurate predictions of discharge timing and in-hospital mortality could improve hospital efficiency, but clinician estimates are often inconsistent and imprecise. We evaluated if machine learning models could concurrently predict in-hospital mortality and length of stay (LoS) more reliably.

**Methods:** We used electronic healthcare data from 01-November-2021 to 31-October-2024 from Oxfordshire, UK, using two years of data for training and evaluating models using the final year’s data. The performance of task-specific extreme gradient boosting (XGB), logistic regression (LR), and multilayer-perceptron (MLP) models for the two tasks: (i) mortality prediction and (ii) LoS prediction, were compared with that of a single multiclass XGB model predicting combinations of LoS and mortality, and an MLP-based multi-task learning model predicting both outcomes simultaneously. Predictions from the best-performing models were compared to discharge predictions made by clinicians.

**Findings:** Clinicians provided relevant discharge predictions for only 3-5% of admissions, mostly close to discharge. Task-specific XGB models achieved an area under the receiver operating curve of 0.92 and 0.92 for predicting mortality, and 0.83 and 0.72 for predicting LoS quartiles, in elective and emergency admissions respectively, outperforming task-specific LR and MLP models. Neither the multiclass XGB nor the MLP-based multi-task models, predicting both outcomes simultaneously, consistently improved performance. The best-performing task-specific XGB models matched clinician LoS prediction accuracy in elective admissions, and significantly outperformed clinicians in emergency admissions (p<0.001).

**Interpretation:** Machine learning models can predict in-hospital mortality and LoS as well or better than clinicians and have potential to enhance discharge planning and hospital resource management.

**Funding:** National Institute for Health Research (NIHR) Biomedical Research Centre, Oxford, and NIHR Health Protection Research Unit in Healthcare Associated Infections and Antimicrobial Resistance at Oxford University in partnership with the UK Health Security Agency (UKHSA).

## Introduction

Increasing demand for healthcare services has placed significant pressure on limited available resources, highlighting the need to enhance hospital efficiency and capacity management^1^. Discharge planning is a critical component of operational management, and early discharge planning helps reduce length of stay (LOS) and readmissions. Some hospitals have implemented discharge planning interventions, asking healthcare professionals to predict the date of discharge^2–4^. However, there is a lack of evidence examining the accuracy of these predictions, which may also be resource intensive to generate and inconsistently performed^5^. Identifying patients at risk of in-hospital mortality is important to bed-state predictions and also has potential to improve patient outcomes.

Several machine learning models using electronic healthcare (EHR) data have been developed to predict either mortality^6–10^ or LoS^11–15^ among hospitalised patients, with modest performance. However, these studies typically focus on a single prediction task, overlooking the interconnected nature of real-world clinical outcomes. Both discharge and mortality may lead to a bed becoming available, but these events are preceded by different health trajectories, with models having to track recovery or deterioration respectively. A few studies have explored predicting both mortality and LoS using deep learning models, but these tasks were trained separately^16,17^. Multi-task learning, a technique that enables simultaneous prediction of multiple related outcomes by sharing feature representations between tasks^18^, offers a promising alternative. The approach has shown success in various healthcare applications, e.g. cancer prognosis^19^, postoperative complications^20^, and Alzheimer’s disease diagnosis^21^. One study used a neural network-based multi-task learning model to predict inpatient flow and LoS with improved accuracy compared to single-task models, although it did not address mortality^22^. A recent study introduced a multi-modal multi-task learning framework to perform LoS regression and mortality classification tasks simultaneously using the Medical Information Mart for Intensive Care database, achieving better performance than conventional methods^23^. However, there was limited evaluation of practical application of this model, and no comparison made with clinician predictions.

In this study, we first examined the accuracy of clinician-provided discharge estimates from a large UK teaching hospital group. We then built machine learning models using diverse EHR-derived features to predict inpatient mortality and LoS quartiles, comparing the performance and computational efficiency of separate task-specific models for mortality and LoS, with a multi-class model predicting combinations of mortality and LoS together, and a multi-task learning model predicting both mortality and LoS simultaneously, but as separate outcomes. Last, we assessed the models’ predictions against those of clinicians to evaluate their practical utility.

## Methods

### Data extraction

We used data from the Infections in Oxfordshire Research Database (IORD), which includes deidentified electronic health records from Oxford University Hospitals (OUH), four teaching hospitals serving around 1% of the UK population. IORD has approvals from the National Research Ethics Service South Central-Oxford C Research Ethics Committee (19/SC/0403), Health Research Authority and Confidentiality Advisory Group (19/CAG/0144) as a deidentified database without individual consent.

We extracted data on all adult inpatients (≥16 years) between 01-November-2021 and 31- October-2024. Admissions were classified as elective (scheduled in advance) or emergency (details in **Supplementary Methods**).

Clinician discharge date predictions were made by individual clinicians or during multi- disciplinary team meetings and recorded in the EHR. Not all patients had predictions documented, as this was not prioritised in all areas of the hospitals.

### Features and preprocessing

To build prediction models, we used the same feature set and preprocessing as previously described^24^. A total of 1,152 features were created and grouped into 15 feature categories based on domain knowledge and previous literature (**Supplementary Methods**), including index date-related features, demographics, comorbidities, current admission, ward stay, current diagnostic category, procedures, antibiotics prescriptions, medication, microbiology tests, radiology investigation, readmissions and previous hospital stay, hospital capacity factors reflecting crowdedness, vital signs, and laboratory tests. Missing categorical features were imputed using the mode, while numerical features were imputed using the median.

### Prediction outcome

We predicted in-hospital mortality and future hospital LoS at an index date and time for all patients currently in the hospital. The in-hospital mortality was extracted as a binary event and future hospital LoS were categorised into quartiles, reflecting clinically relevant durations of future stay:

- Elective admissions: Q1: 0-1, Q2: 2-3, Q3: 4-10, Q4: >10 days.
- Emergency admissions: Q1: 0-2, Q2: 3-6, Q3: 7-14, Q4: >14 days.

Predictions were made daily at 12 noon to align with the end of typical hospital ward rounds. Each patient contributed once to the dataset each day during their hospital admission.

Separate models were built for emergency and elective admissions as predictive features plausibly differ by the admission type. Models were trained using data from the first two years (01-November-2021 to 31-October-2023) and evaluated using data from the final year (01-November-2023 to 31-October-2024).

### Model architectures

In our single-task model analysis, we used binary extreme gradient boosting (XGB) to predict mortality, and a separate multiclass XGB to predict hospital LoS (see **Supplementary Methods** for details of hyperparameter tuning). We used 20% of the training dataset as validation data for feature selection (retaining the 200 most informative features based on this optimising performance in a previous study^24^), calibrating the predicted probability from XGB models using isotonic regression, and determining the best threshold for predicting mortality by optimising the F1 score (jointly maximising precision and recall). We compared the XGB models to logistic regression (LR) and multilayer perceptron (MLP) models, using the same 200 features selected for the XGB models (details in **Supplementary Methods**).

We next fitted an XGB model similarly to above but combining mortality and LoS quartiles into a single multiclass model with seven outcome categories comprising linked mortality and LoS outcomes:

- Elective: died in hospital within 0-1 days, 2-3 days, or 4-10 days; discharged alive within 0-1 days (Q1), 2-3 days (Q2), or 4-10 days (Q3); or remained in hospital >10 days (Q4).
- Emergency: died in hospital within 0-2 days, 3-6 days, or 7-14 days; discharged alive within 0-2 days (Q1), 3-6 days (Q2), or 7-14 days (Q3); or remained in hospital >14 days (Q4).

Finally we fitted a multi-task learning model^18,25^ using an MLP architecture based on the same feature set as the single-task models to predict mortality and LoS quartiles simultaneously. The same loss functions and hyperparameters were used as in the single-task models. To properly combine the task-specific losses, we compared different loss weighting strategies, including uniform weighting (with different weights), random loss weighting^26^, and uncertainty weighting^27^.

### Performance assessment

Model predictions were evaluated using established metrics, comparing performance to actual events. We also compared the performance of the LoS models with clinician predictions. As clinician predictions were made at random timepoints throughout the day, for comparisons a separate model prediction was made, using the time of the prediction, rather than 12 noon, as the index date and time for the data extract.

## Results

### Patient characteristics

From 01-November-2021 to 31-October-2024, a total of 37,574 adult elective admissions (31,306 patients) and 154,849 emergency admissions (94,702 patients) were included in the analyses. The median age was 65 (IQR: 46-79) years, 98,222 (51.0%) patients were female, and 136,670 (71.0%) were recorded as white ethnicity (ethnicity missing in 43,214, 22.5%).

Mortality rates were 0.4% (158/37,574) for elective and 4.3% (6629/154,849) for emergency admissions. The future LoS followed a skewed distribution, with medians of 2.1 (IQR: 1.2-4.6) days for elective and 2.2 (0.6-7.0) days for emergency admissions (**Figure S1**). Demographic characteristics were similar between the training and test datasets (**Table 1**).

**Table 1.**
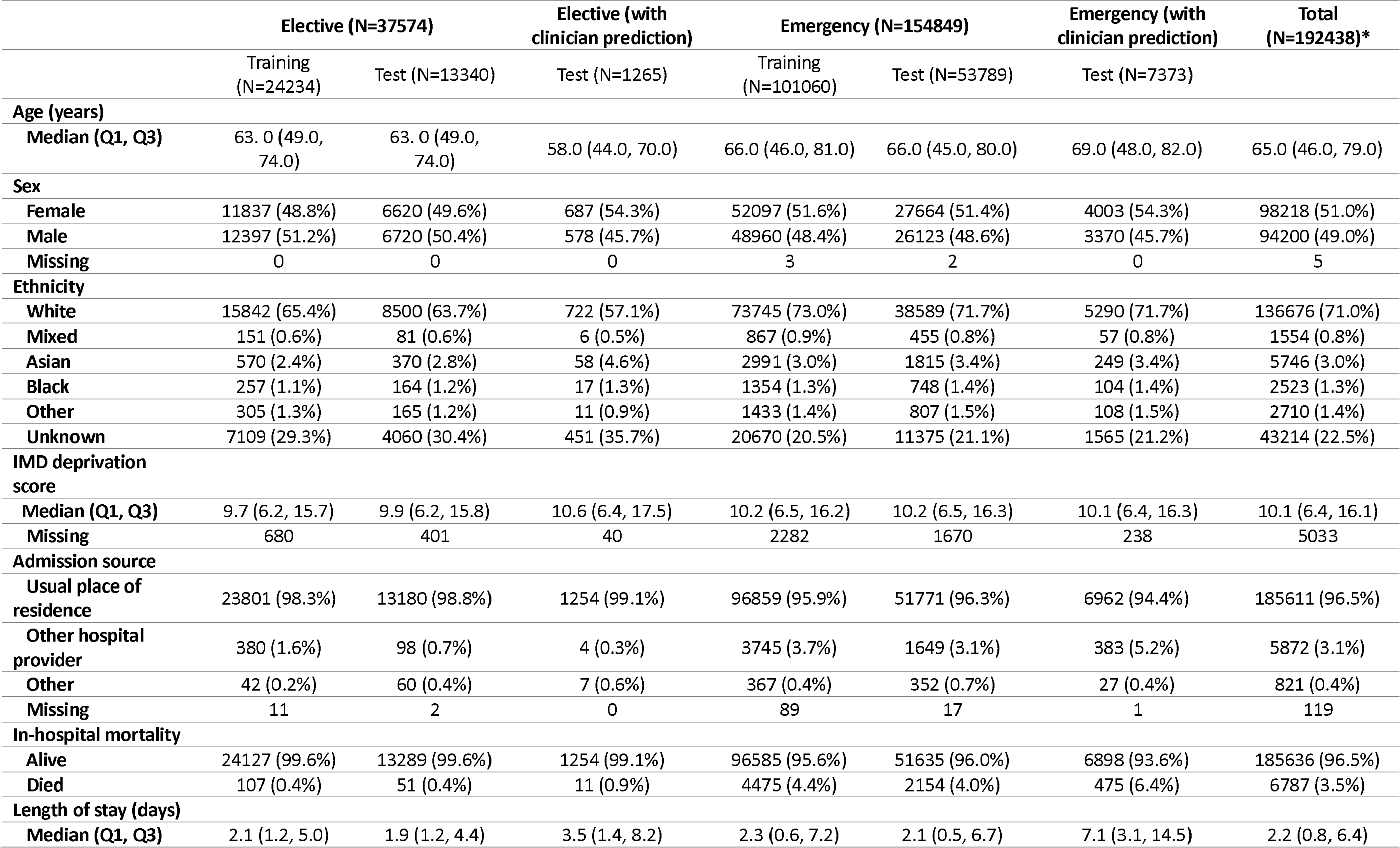
Baseline characteristics of 37,574 elective and 154,849 emergency admissions between 01 November 2021 to 31 October 2024 used in training and testing discharge prediction models. Characteristics of 1,265 elective admissions and 7,373 emergency admissions with clinician predictions between 01 November 2023 to 31 October 2024 are presented. *Admissions with clinician predictions were not included as overlapped with test data. IMD: index of multiple deprivation (higher scores indicate greater deprivation, range 0-77).

|  | Elective (N=37574) |  | Elective (with<br>clinician prediction) | Emergency (N=154849) |  | Emergency (with<br>clinician prediction) | Total<br>(N=192438)* |
| --- | --- | --- | --- | --- | --- | --- | --- |
|  | Training<br>(N=24234) | Test (N=13340) | Test (N=1265) | Training<br>(N=101060) | Test (N=53789) | Test (N=7373) |  |
| **Age (years)** |  |  |  |  |  |  |  |
| Median (Q1, Q3) | 63.0 (49.0, 74.0) | 63.0 (49.0, 74.0) | 58.0 (44.0, 70.0) | 66.0 (46.0, 81.0) | 66.0 (45.0, 80.0) | 69.0 (48.0, 82.0) | 65.0 (46.0, 79.0) |
| **Sex** |  |  |  |  |  |  |  |
| Female | 11837 (48.8%) | 6620 (49.6%) | 687 (54.3%) | 52097 (51.6%) | 27664 (51.4%) | 4003 (54.3%) | 98218 (51.0%) |
| Male | 12397 (51.2%) | 6720 (50.4%) | 578 (45.7%) | 48960 (48.4%) | 26123 (48.6%) | 3370 (45.7%) | 94200 (49.0%) |
| Missing | 0 | 0 | 0 | 3 | 2 | 0 | 5 |
| **Ethnicity** |  |  |  |  |  |  |  |
| White | 15842 (65.4%) | 8500 (63.7%) | 722 (57.1%) | 73745 (73.0%) | 38589 (71.7%) | 5290 (71.7%) | 136676 (71.0%) |
| Mixed | 151 (0.6%) | 81 (0.6%) | 6 (0.5%) | 867 (0.9%) | 455 (0.8%) | 57 (0.8%) | 1554 (0.8%) |
| Asian | 570 (2.4%) | 370 (2.8%) | 58 (4.6%) | 2991 (3.0%) | 1815 (3.4%) | 249 (3.4%) | 5746 (3.0%) |
| Black | 257 (1.1%) | 164 (1.2%) | 17 (1.3%) | 1354 (1.3%) | 748 (1.4%) | 104 (1.4%) | 2523 (1.3%) |
| Other | 305 (1.3%) | 165 (1.2%) | 11 (0.9%) | 1433 (1.4%) | 807 (1.5%) | 108 (1.5%) | 2710 (1.4%) |
| Unknown | 7109 (29.3%) | 4060 (30.4%) | 451 (35.7%) | 20670 (20.5%) | 11375 (21.1%) | 1565 (21.2%) | 43214 (22.5%) |
| **IMD deprivation score** |  |  |  |  |  |  |  |
| Median (Q1, Q3) | 9.7 (6.2, 15.7) | 9.9 (6.2, 15.8) | 10.6 (6.4, 17.5) | 10.2 (6.5, 16.2) | 10.2 (6.5, 16.3) | 10.1 (6.4, 16.3) | 10.1 (6.4, 16.1) |
| Missing | 680 | 401 | 40 | 2282 | 1670 | 238 | 5033 |
| **Admission source** |  |  |  |  |  |  |  |
| Usual place of residence | 23801 (98.3%) | 13180 (98.8%) | 1254 (99.1%) | 96859 (95.9%) | 51771 (96.3%) | 6962 (94.4%) | 185611 (96.5%) |
| Other hospital provider | 380 (1.6%) | 98 (0.7%) | 4 (0.3%) | 3745 (3.7%) | 1649 (3.1%) | 383 (5.2%) | 5872 (3.1%) |
| Other | 42 (0.2%) | 60 (0.4%) | 7 (0.6%) | 367 (0.4%) | 352 (0.7%) | 27 (0.4%) | 821 (0.4%) |
| Missing | 11 | 2 | 0 | 89 | 17 | 1 | 119 |
| **In-hospital mortality** |  |  |  |  |  |  |  |
| Alive | 24127 (99.6%) | 13289 (99.6%) | 1254 (99.1%) | 96585 (95.6%) | 51635 (96.0%) | 6898 (93.6%) | 185636 (96.5%) |
| Died | 107 (0.4%) | 51 (0.4%) | 11 (0.9%) | 4475 (4.4%) | 2154 (4.0%) | 475 (6.4%) | 6787 (3.5%) |
| **Length of stay (days)** |  |  |  |  |  |  |  |
| Median (Q1, Q3) | 2.1 (1.2, 5.0) | 1.9 (1.2, 4.4) | 3.5 (1.4, 8.2) | 2.3 (0.6, 7.2) | 2.1 (0.5, 6.7) | 7.1 (3.1, 14.5) | 2.2 (0.8, 6.4) |

### Clinician predictions

During the test period, clinicians made a total of 2,117 and 14,334 predictions during 1,265 (3.4%) elective and 7,363 (4.8%) emergency admissions, respectively. Relevant predictions were made on 2,081 (98.3%) and 13,946 (97.3%) days (the remainder had an anticipated discharge date earlier than the date that the prediction was made). In elective admissions, most clinician predictions were made in a surgical subspecialty (41.6%) or a medical subspecialty (36.3%). In emergency admissions, most predictions were made in acute, emergency and geriatric medicine (35.6%), followed by a medical subspecialty (27.9%), and acute and general surgery (16.1%). The median LoS for patients with clinician predictions was longer than in the overall dataset, being 3.5 (1.4-8.2) days for elective and 7.1 (3.1-14.5) days for emergency admissions (**Table 1**).

Most clinician predictions were made in the morning between 9-11am for elective and 7- 11am for emergency admissions (**Figure 1 a,b**). Predictions were typically made 1.2 (IQR 0.4- 3.6) days and 1.9 (0.7-4.3) days from admission (**Figure 1 c,d**), and 1.4 (0.3-6.2) days and 4.3 (1.3-11.2) days before actual discharge, for elective and emergency admissions, respectively (**Figure 1 e,f**). The prediction horizon was short; clinicians tended to predict patients being discharged in 1.5 (0.5-4.5) days and 2.6 (0.6-3.7) days for elective and emergency admissions, respectively. 981 (47.1%) elective and 3,744 (26.8%) emergency predictions were for discharge that day or the following day.

**Figure 1.**
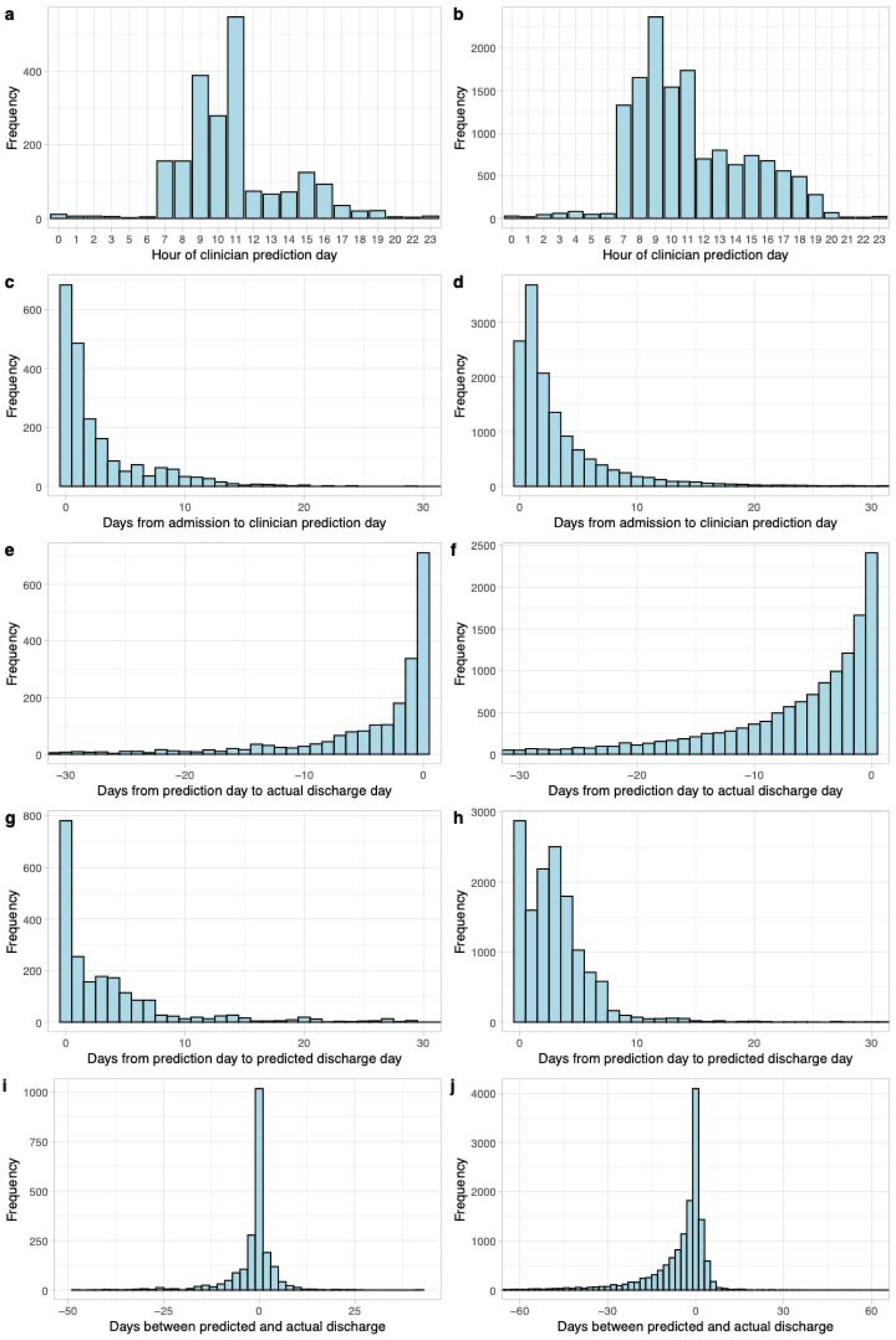
Clinician predictions of discharge. a, b) Time of the day of clinician prediction; c, d) Days from admission to when clinicians made predictions (prediction day); e, f) Days from prediction day to actual discharge day; g, h) Days from prediction day to predicted discharge day; I, j) Days between clinician prediction and actual discharge. a, c, e, g, i are elective admissions, b, d, f, h, j are emergency admissions.

For elective admissions, 859 (41.2%) predictions were correctly made, 661 (31.8%) were earlier than the actual discharge date, and 561 (27.0%) were later than the actual discharge date. For emergency admissions, 3,038 (21.2%) were correctly made, 7,518 (53.9%) were earlier than the actual discharge date, i.e., tended to be overly optimistic, and 3,390 (24.3%) were later than the actual discharge date (**Figure 1 g,h**). Overall, clinician predictions were made inconsistently and had suboptimal accuracy.

### Single-task model performance

Predicting discharge at 12 noon, 186,627 and 960,354 patient-days were included in the analyses for elective and emergency admissions respectively.

#### Mortality prediction

For elective admissions, the XGB model achieved an area under the receiver operating curve (AUROC) of 0.917 (95%CI 0.910-0.924) and an area under the precision recall curve (AUPRC) of 0.219 (0.146-0.268) for predicting inpatient mortality. Using a probability threshold that optimised F1 score in the validation dataset, the positive predictive value (PPV) was 0.262, negative predictive value (NPV) 0.989, and F1 score 0.242 (**Table S1**). The LR model yielded a lower AUROC of 0.876 (0.865-0.888) and an AUPRC of 0.204 (0.187-0.223), while the MLP model achieved a similar AUROC of 0.922 (0.916-0.929) and an AUPRC of 0.206 (0.181-0.227) (**Figure 2 a,b**).

**Figure 2.**
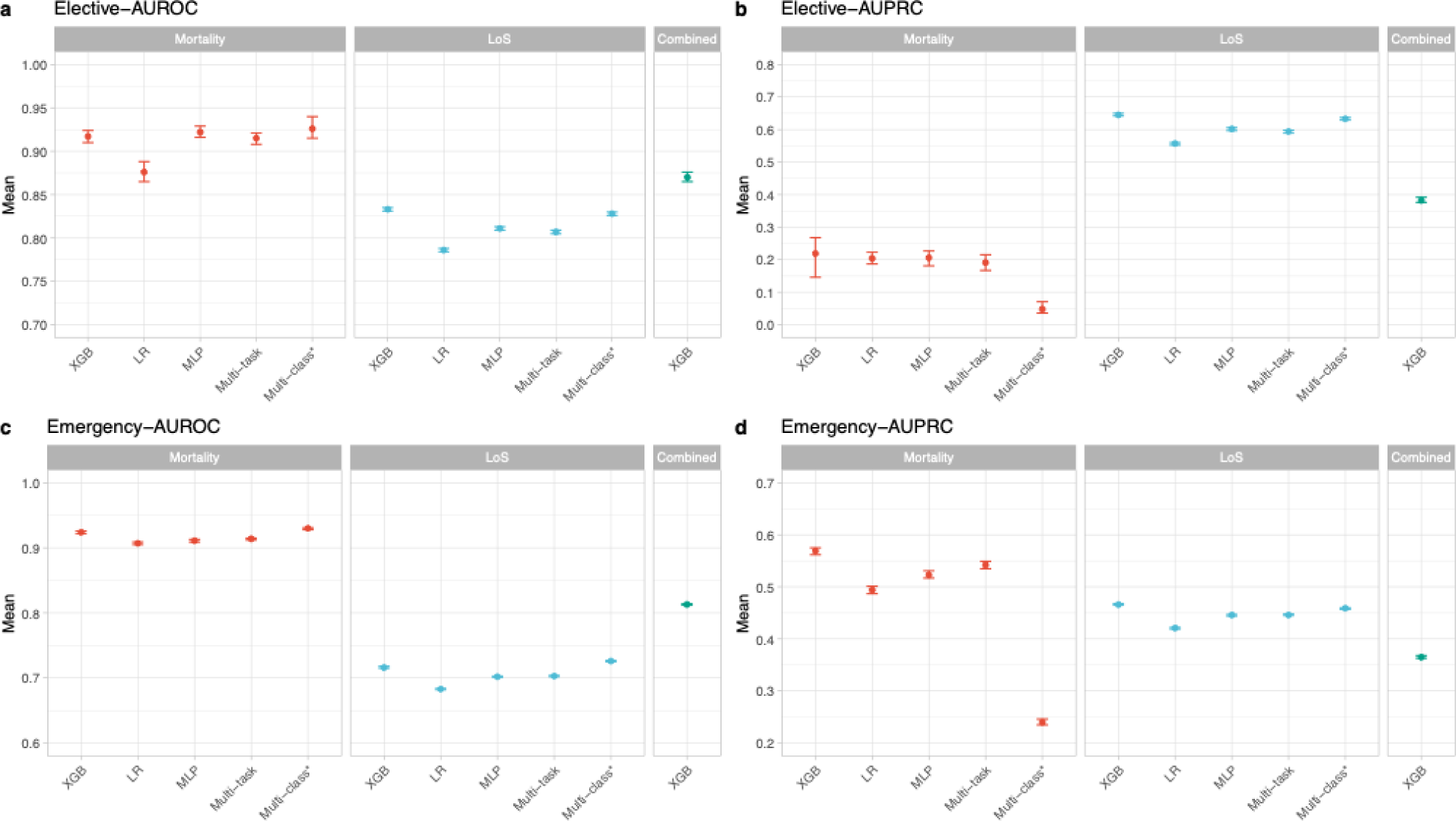
Comparison of model performance. Extreme gradient boosting (XGB) model, logistic regression (LR) model, multi-layer perceptron (MLP) model, and multi-task learning model were built for predicting mortality and future length of stay (LoS), and multiclass XGB was built for predicting combined mortality and LoS.*Macro-averaged performance for mortality classes (died in hospital) or LoS classes (Q1-Q4) from the multiclass XGB model predicting combined mortality and LoS. a) Area under the receiver-operating curve (AUROC) for elective admissions; b) Area under the precision-recall curve (AUPRC) for elective admissions; c) Area under the receiver-operating curve (AUROC) for emergency admissions; d) Area under the precision-recall curve (AUPRC) for emergency admissions. Mean with 95% confidence intervals are calculated using bootstrap. In-hospital mortality occurred following 2,982 (1.6%) elective and 79,646 (8.3%) emergency patient-days. Across all patient-days studied, the median future LoS was 3.2 (IQR 1.1-10.0) days and 5.9 (1.9-14.2) days for elective and emergency admissions, respectively (i.e. longer than the overall LoS as longer admissions contributed proportionally more to the dataset than shorter admissions). In elective admissions, patient-days were distributed as 0-1 day (Q1, 20.1%), 2-3 days (Q2, 25.9%), 4-10 days (Q3, 29.0%), and >10 days (Q4, 25.0%). In emergency admissions, patient-days were distributed as 0-2 days (Q1, 25.8%), 3-6 days (Q2, 24.8%), 7-14 days (Q3, 23.8%), and >14 days (Q4, 25.6%).

For emergency admissions, the XGB model achieved a similar AUROC at 0.924 (0.922-0.926), and a higher AUPRC of 0.569 (0.562-0.575) reflecting the higher mortality rate. At the optimal F1 threshold, the PPV was 0.532, NPV 0.963, and F1 score 0.551 (**Table S1**). The XGB was significantly better than the LR model (AUROC: 0.907 (0.905-0.909); AUPRC: 0.494 (0.487-0.501)), and also better than the MLP model (AUROC: 0.911 (0.909-0.913); AUPRC: 0.523 (0.517-0.531)) (**Figure 2 c,d**, **Table S2**).

For both elective and emergency admissions, AUROC remained consistent with varying time since admission, while AUPRC increased due to a higher mortality rate with longer LoS. Performance was higher when a prediction was made closer to discharge (**Figure 3**).

**Figure 3.**
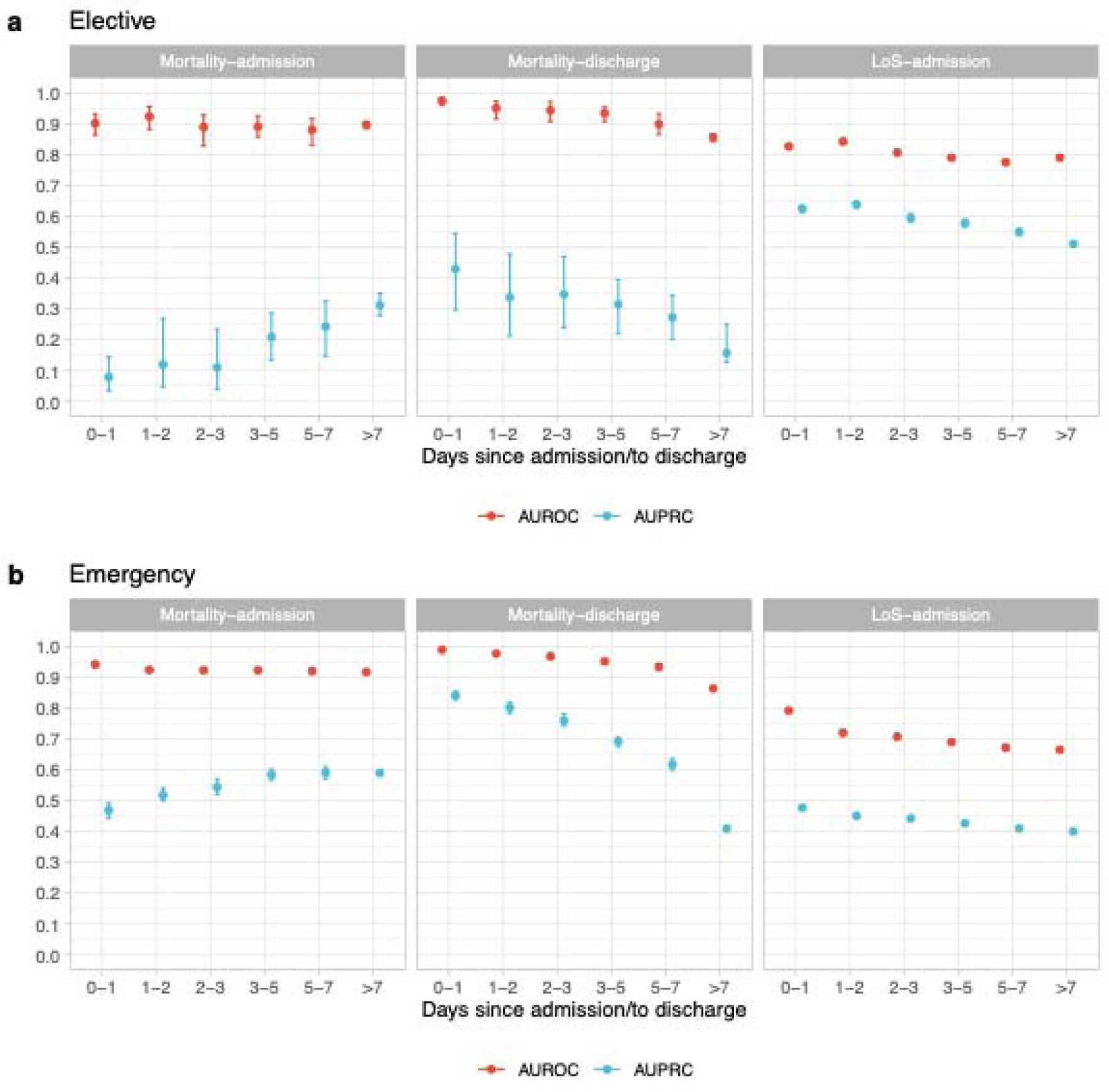
Subgroup performance by time since admission (for predicting both mortality and length of stay) or time to discharge (for predicting mortality) in the test data using extreme gradient boosting model. a) elective admissions; b) emergency admissions. Mean with 95% confidence intervals are calculated using bootstrap.

#### Future LoS prediction

The XGB model achieved a macro-averaged (giving the same weight to each LoS category or class) AUROC of 0.833 (0.831-0.835), an AUPRC of 0.646 (0.642-0.650), and an F1 score of 0.587 (0.584-0.591) (**Table S1**). Q4 had the highest individual one-vs-rest AUROC (0.901), followed by Q1 (0.876), Q2 (0.788), and Q3 (0.768), i.e. patients with the longest or shortest LoS were simplest to identify (**Figure S2**). LR and MLP models performed slightly worse, with macro-averaged AUROCs of 0.786 (0.784-0.788) and 0.811 (0.809-0.813), respectively (**Figure 2**).

Performance was generally lower for emergency admissions compared to elective admissions, with the XGB model achieving a macro-averaged AUROC of 0.716 (0.715-0.718) and an AUPRC of 0.466 (0.465-0.468), and the highest individual AUROC in Q1 (0.813) and Q4 (0.760) **(Figure S2**). LR and MLP models showed lower performance with AUROCs of 0.683 (0.682-0.684) and 0.702 (0.701-0.703), respectively (**Figure 2, Table S1**).

AUROC was relatively stable by time since admission (around 0.80) for elective admissions, but decreased from 0.80 the day after admission to 0.65 >7 days since admission for emergency admissions, reflecting the difficulty in predicting discharge for patients with longer hospital stays (**Figure 3**).

#### Feature importance

The top 20 features ranked by importance are shown in **Figure 4**. The most predictive features for in-hospital mortality were Elixhauser and Charlson comorbidity scores, which were selected as the top 5 most important features for both elective and emergency admissions. Other important features were the number of specialties involved in the current admission to date and various laboratory test measurements (activated partial thromboplastin time, urea, haemoglobin, lymphocytes, lactate) for elective admissions, and age, historical LoS, oral medications, albumin measurement, and body mass index for emergency admissions.

**Figure 4.**
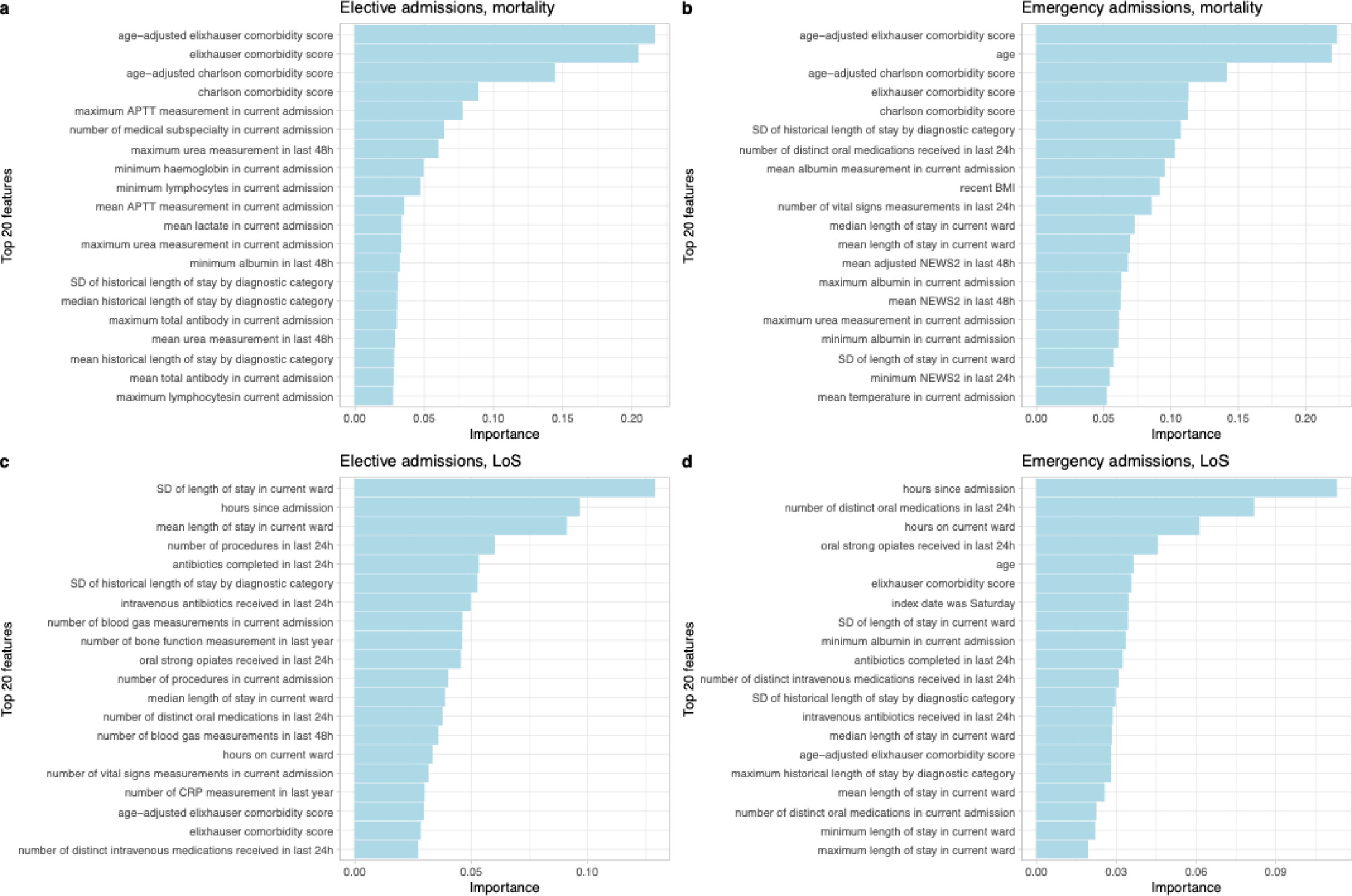
Feature importance from the extreme gradient boosting models predicting in-hospital mortality and future hospital length of stay (LoS) using SHapley Additive exPlanations values for elective and emergency admissions. The top 20 most predictive features are shown in the order of predictiveness. APTT: activated partial thromboplastin time; SD: standard deviation; BMI: body mass index; CRP: C-reactive protein.

Hours since admission, LoS on current ward, antibiotics, and medications were selected as the most important features predicting future LoS for both elective and emergency admissions.

### Combined multi-class model performance

Using a multi-class XGB model to predict combined mortality and LoS outcomes (**Table S3**), elective admissions had a macro-averaged AUROC of 0.870 (95%CI 0.865-0.876), with those dying in hospital 0-1 days later having the highest AUROC (0.960), followed by deaths in hospital 2-3 days later (0.924) and remaining in hospital for >10 days longer (0.898). The macro-averaged AUPRC was 0.381 (0.376-0.392), with those dying in hospital in the next 0- 10 days having low values <0.1 due to the low prevalence (**Table S2, Figure S3**). Macro-averaging the three mortality classes resulted in an AUROC of 0.926 (0.915-0.940) (cf. separate binary XGB mortality model: 0.917 (0.910-0.924), and LoS classes (discharged Q1- Q4) 0.828 (0.826-0.830) (cf. separate XGB LoS model: 0.833 (0.831-0.835) (**Figure 2**).

For emergency admissions, the macro-averaged AUROC was 0.813 (0.812-0.814) and the AUPRC 0.365 (0.363-0.368), with the highest AUROC observed for admissions with mortality outcomes (deaths in 0-2, 3-6 and 7-14 days: 0.966, 0.929, 0.894) (**Table S1, Figure S3**).

Macro-averaging mortality classes resulted in an AUROC of 0.930 (0.928-0.931) (cf. separate model: 0.924 (0.922-0.926)), and LoS classes 0.726 (0.725-0.727) (cf. separate model 0.716 (0.715-0.718)) (**Figure 2**).

### Multi-task model performance

We trained a multi-task MLP model using the same feature set selected from single-task XGB models. Comparing different weighting strategies, the performance for mortality prediction was relatively stable, while higher performance was achieved for LoS prediction when a higher weight was assigned (**Table S4**). Therefore, the final models used a uniform weight of 1:10 for mortality:LoS.

For elective admissions, the multi-task model achieved an AUROC of 0.915 (0.908-0.921) for mortality (vs. 0.917 (95%CI 0.910-0.924) in separate XBG models) and 0.807 (0.805-0.813) for LoS prediction in the test data (vs. 0.833 (0.831-0.835)). For emergency admissions, AUROCs were 0.914 (0.912-0.915) for mortality (vs. 0.924 (0.922-0.926)) and 0.703 (0.702-0.704) for LoS (vs. 0.716 (0.715-0.718)). Hence, while mortality performance was comparable to single-task XGB or MLP models, LoS performance was lower than XGB but comparable to MLP (**Table S1, Figure 2**).

### Model training times

To compare training times across models, we calculated the total time for hyperparameter tuning, model fitting, and prediction. To ensure a fair comparison, 30 optimisation trials were used for each model, and the terminator improvement plot was examined to ensure the number of trials was sufficient. For elective admissions, there was a time saving associated with the combined models, the training time for the multi-task MLP model and the multi-class XBG model was shorter than the combined training time for the separate mortality and LoS single-task XGB or MLP models. For emergency admissions, the multi-task MLP model was faster than the combined mortality and LoS single-task MLP models but was slower than the sum of the time taken for the two single-task XGB models or for the multi-class XGB model (**Figure 5**).

**Figure 5.**
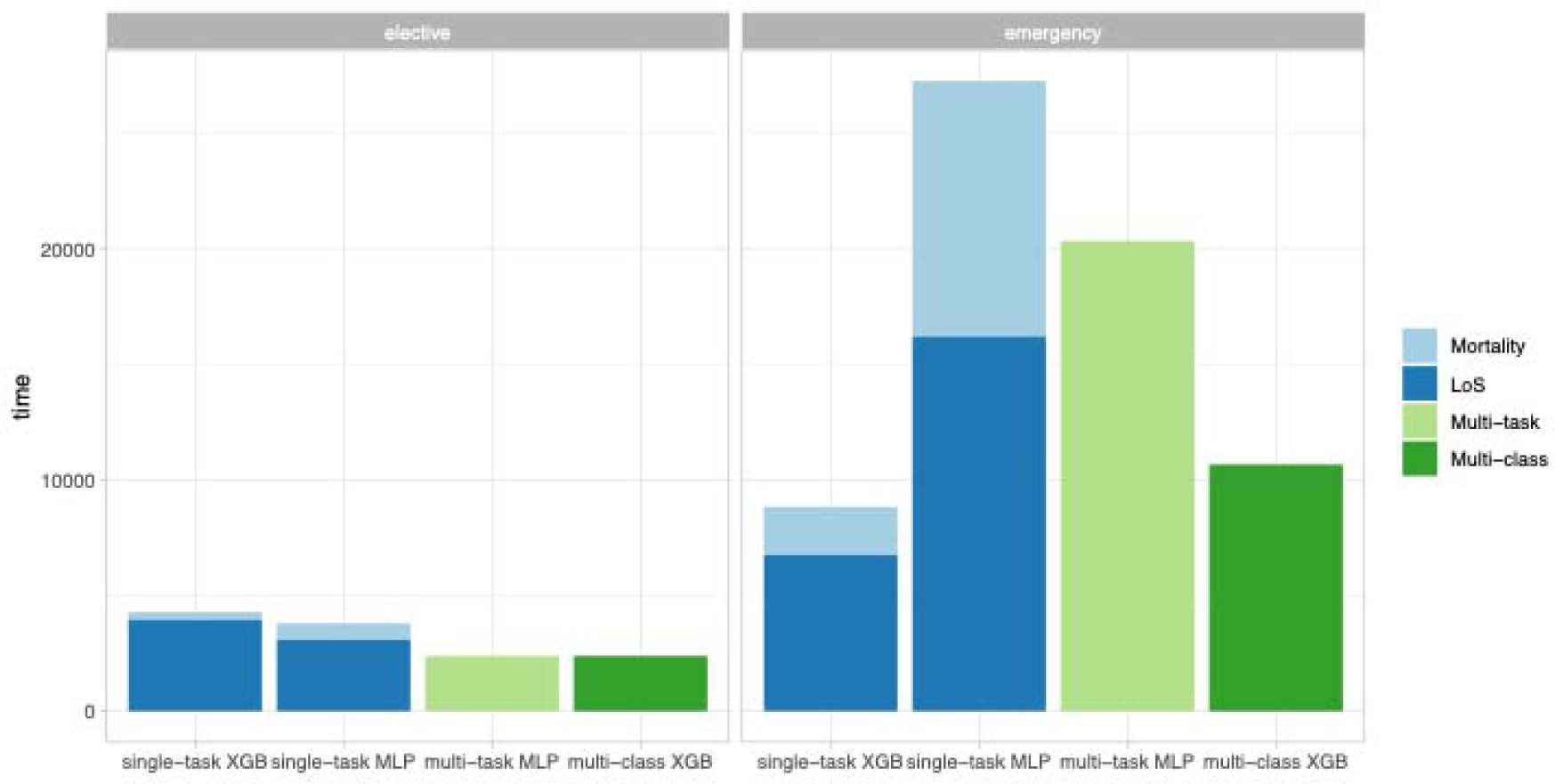
Comparison of training time for single-task extreme gradient boosting (XGB) model, single-task multilayer perceptron (MLP), multi-task MLP, and combined multi-class XGB.

### Comparison with clinician predictions

Using the best-performing single-task XGB model to predict future LoS at the same time point as clinicians had made a prediction, the model performance vs. the actual discharge date resulted in a macro-averaged AUROC and AUPRC of 0.826 and 0.648 for elective admissions, and 0.711 and 0.492 for emergency admissions. The model achieved similar accuracy to clinicians for elective admissions (0.566 vs. 0.585, p=0.14 using McNemar’s test), but significantly outperformed clinicians for emergency admissions (0.442 vs. 0.402, p<0.001).

The macro-averaged F1 scores were 0.550 (model) vs. 0.538 (clinicians) for elective and 0.422 (model) vs. 0.308 (clinicians) for emergency admissions (**Figure 6 a,b**). The model was better at predicting those who were more likely to stay longer, reflecting that clinicians tended to be overly optimistic. For example, clinicians correctly predicted 70.0% and 56.0% of patients who were discharged in 0-2 or 3-6 days, vs. 64.5% and 28.8% from the model, while clinicians only correctly predicted 13.6% and 2.2% of patients who were discharged in 7-14 and >14 days, vs. 35.8% and 36.7% from the model (**Figure 6 c-f**).

**Figure 6.**
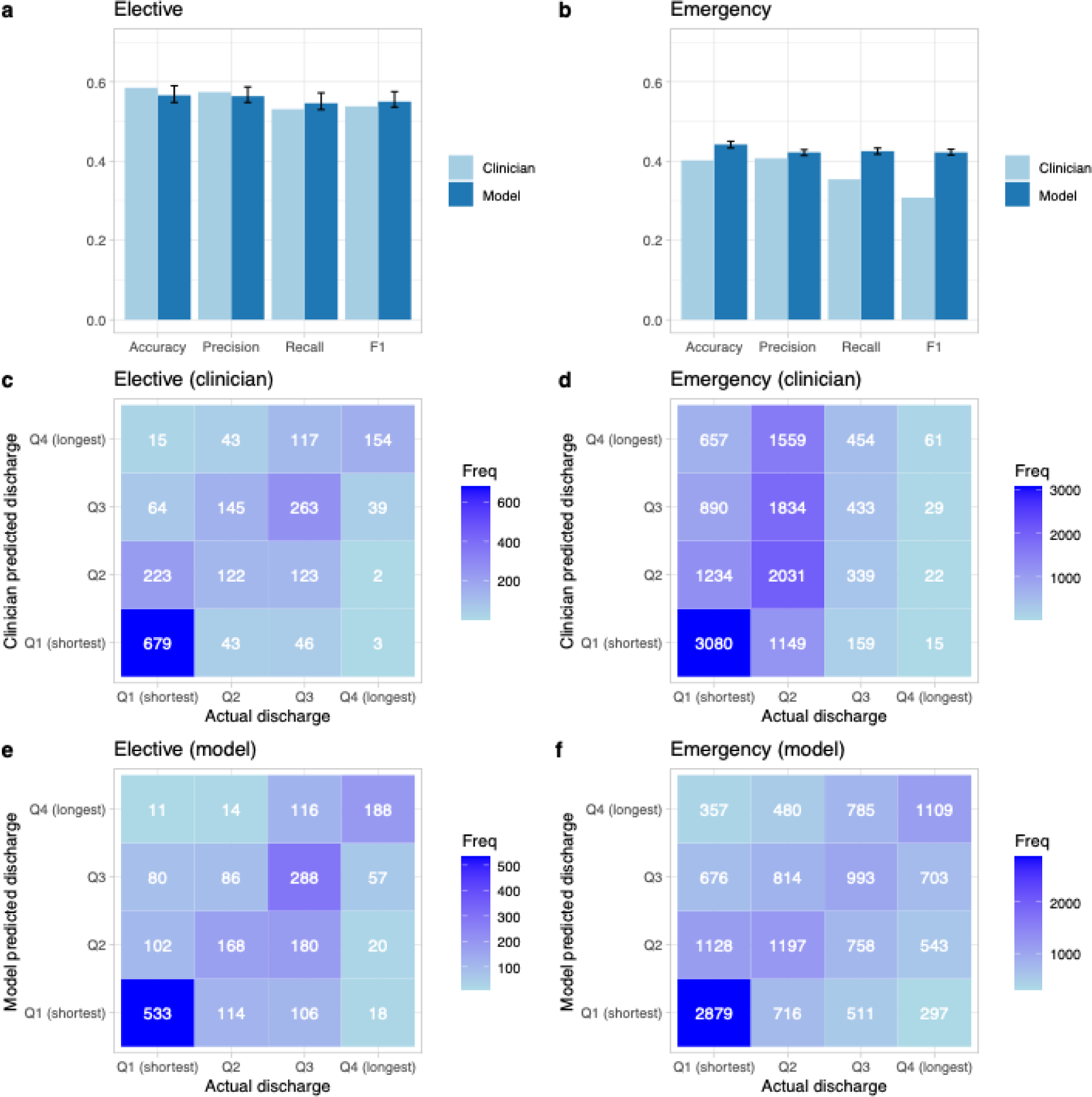
Comparison between clinician predictions and model predictions on future hospital length of stay (LoS) in the test dataset where clinicians made predictions. a) Comparison of accuracy, precision, recall, and F1 score between clinician and the best-performing extreme gradient boosting (XGB) model for elective admissions. b) Comparison of accuracy, precision, recall, and F1 score between clinician and the best-performing extreme gradient boosting (XGB) model for emergency admissions. c) Confusion matrix for clinician predicted and actual LoS quartiles for elective admissions. d) Confusion matrix for clinician predicted and actual LoS quartiles for emergency admissions. e) Confusion matrix for model predicted and actual LoS quartiles for elective admissions. f) Confusion matrix for model predicted and actual LoS quartiles for emergency admissions. Q1: below 25% quantile (0-1 days for elective, 0-2 for emergency); Q2: 25% to 50% quantile (2-3 days for elective, 3-6 days for emergency); Q3: 50% to 75% quantile (4-10 days for elective, 7-14 days for emergency); Q4: above 75% quantile (>10 days for elective, >14 days for emergency). Mean with 95% confidence intervals are calculated using bootstrap.

## Discussion

Exploiting a wide range of features in EHRs, we developed machine learning models based on XGB and MLP architectures to predict inpatient mortality and future LoS and explored whether a multi-task learning strategy could further improve performance by predicting both outcomes simultaneously. Our models for predicting in-hospital mortality demonstrated strong predictive performance, with an AUROC of 0.92 for both elective and emergency admissions. For LoS prediction in clinically relevant quartiles, the best models achieved an AUROC of 0.83 and 0.72 for elective and emergency admissions, respectively. Notably, our models outperformed clinicians in predicting LoS for emergency patients, whilst providing similar performance in elective patients.

Various machine learning approaches have been used previously to predict in-hospital mortality among patients admitted as emergencies, with AUROCs ranging from 0.80 to 0.92^7–9,28–30^. Our model is amongst the best reported for this task, and performance was stable when predictions were made at different times from admission. We achieved similar AUROC for both elective and emergency admissions.

Building on our earlier work predicting imminent hospital discharge within 24 hours, this study focused on longer LoS. Unlike most studies treating LoS as a continuous outcome^11,23^ or predicting prolonged LoS as a binary outcome^15,31^, we categorised LoS into quartiles to provide more granular clinically-relevant predictions for resource management. This approach aligns with recent research that reported AUROC values of 0.664 using structured data and 0.787 with a Large Language Model (LLM)^32^. We achieved better performance than their model using structured data for both admission types (AUROC: 0.833 (elective) and 0.716 (emergency)), and better performance than the LLM for elective admissions, potentially because we included much more detailed features. Our predictions were more accurate for elective than emergency admissions, reflecting more uncertainty in emergency admissions and greater requirements for social care post-discharge. We achieved better performance than a previous study predicting multiclass-LoS for planned admissions^13^ (AUPRC 0.646 vs. 0.556).

Comorbidity scores were the strongest predictors of mortality for both admission types. For elective admissions, laboratory test measurements were also critical, whereas emergency predictions relied more on age, body mass index, historical LoS, and vital signs. For LoS prediction, the most influential features included hours since admission, hospital capacity factors, non-antibiotic medications, and antibiotic use, reflecting clinical improvement and operational pressures. Age was a top predictor in emergency but not elective admissions.

Combined multi-class and multi-task models potentially improve performance and reduce training time. However, overall, the performance improvements we observed were inconsistent and only modest when present. The multiclass model marginally improved LoS predictions for emergency admissions (AUROC: 0.726 [95%CI 0.725-0.727] vs. 0.716 [0.715- 0.718]) but not for elective admissions. The multi-task model did not outperform the single-task MLP or XGB models, particularly for predicting LoS, possibly reflecting XGB’s superior handling of tabular data^33,34^. Similar results have been reported in other multi-class classification problems^35,36^ and also for a multi-task model that did not achieve better performance than individual models predicting mortality, acute kidney injury, and reintubation^20^. Although multi-task training reduced training time for MLP models, XGB models trained within acceptable time periods and demonstrated better predictive accuracy, making them more suitable for clinical applications.

Our models matched or outperformed clinicians in predicting LoS, particularly for emergency admissions. Clinicians tended to overestimate early discharges, while our models provided more accurate predictions for longer stays. Furthermore, clinicians only predicted discharge dates sporadically in our setting (<5% of admissions), and even made non-sensical predictions of discharge dates in the past. Therefore, as well as potentially outperforming clinicians, our models also have the potential to deliver predictions consistently for all patients, and at different timepoints throughout a hospital stay. Our results align with several previous studies showing similar or better performance from models than clinicians in predicting hospital discharges^12,37^. Integrating such automated prediction models into clinical practice remains underexplored and needs further investigation. Using comprehensive data to predict both mortality and LoS, we envision our models could be applied to improve the efficiency of care delivery and patient recovery, and optimal use of healthcare resources.

Limitations of our study include that we did not consider multimodal data such as time-series, free text, or radiology data. Using these data might improve performance but would require more complex model architectures, improved data quality, and additional computational resources. We used data from the same hospital group for training and testing. Future work should perform external validation using data from diverse settings to strengthen the validity and generalisability of findings. We used ICD-10 diagnosis categories as features, these were recorded at discharge, but the information contained within them was likely known to clinicians and could be recorded in real-time. We also lacked data on hospital-level factors such as bed occupancy rates, which could influence predictions, particularly under operational pressure. We did not perform subgroup analyses, but it is possible that refined models for specific patient groups could improve performance, although they would require more resources for model training. Finally, predictions were made at a fixed time daily; however our prior work showed the prediction time of day had limited impact on results^24^.

In conclusion, by integrating machine learning with comprehensive EHR data, our models achieved high performance in predicting in-hospital mortality and future LoS. The LoS model outperformed clinicians for emergency patients, with similar performance for elective patients, but provided predictions on 100% of patient-days in hospital vs just 3-5% with clinicians. While combined multi-class XGB and MLP-based multi-task learning approaches offered some limited performance gains, single-task XGB models generally performed best and could be trained within acceptable time periods, making them well-suited for clinical implementation. Overall, our study demonstrates the potential of machine learning models to effectively manage hospital resources, improve patients’ recovery, and facilitate healthcare delivery.

## Data availability

The datasets analysed during the current study are not publicly available as they contain personal data but are available from the Infections in Oxfordshire Research Database (https://oxfordbrc.nihr.ac.uk/research-themes/modernising-medical-microbiology-and-big-infection-diagnostics/iord-about/), subject to an application and research proposal meeting the ethical and governance requirements of the Database. For further details on how to apply for access to the data and a research proposal template please.

## Code availability

A copy of the analysis code is available at https://github.com/jiaweioxford/ML_mortality_LoS.

## Ethics Committee Approval

Deidentified data were obtained from the Infections in Oxfordshire Research Database which has approvals from the National Research Ethics Service South Central – Oxford C Research Ethics Committee (19/SC/0403), the Health Research Authority and the National Confidentiality Advisory Group (19/CAG/0144), including provision for use of pseudonymised routinely collected data without individual patient consent. Patients who choose to opt out of their data being used in research are not included in the study. The study was carried out in accordance with all relevant guidelines and regulations.

## Acknowledgements

This work uses data provided by patients and collected by the UK’s National Health Service as part of their care and support. We thank all the people of Oxfordshire who contribute to the Infections in Oxfordshire Research Database. Research Database Team: L Butcher, H Boseley, C Crichton, DW Crook, DW Eyre, O Freeman, J Gearing (public representative), R Harrington, K Jeffery, M Landray, A Pal, TEA Peto, TP Quan, J Robinson (public representative), J Sellors, B Shine, AS Walker, D Waller. Patient and Public Panel: M Ahmed, G Blower, J Hopkins, V Lekkos, R Mandunya, S Markham, B Nichols. Members of the panel contribute to setting research priorities and questions, and have reviewed a summary of the data used and of the analytical approach. All results are shared with the panel for comment. DAC was funded by the NIHR Oxford Biomedical Research Centre, an NIHR Research Professorship, and a Royal Academy of Engineering Chair.

## Author Contributions

The study was designed and conceived by AJB, DAC, DWE, and ASW. JW, JG, and DWE curated the data. JW and DWE analysed the data and created the visualisations. JW, ASW, and DWE wrote the first draft of the manuscript. All authors contributed to editing and revising the manuscript.

