## supplementary file for "Concurrent prediction of in-hospital mortality and length of stay using single-task, multi-class, and multi-task machine learning"

### Supplementary Methods

#### Data extraction

We used data from the Infections in Oxfordshire Research Database (IORD), which includes deidentified electronic health records from Oxford University Hospitals (OUH) National Health Service (NHS) Foundation Trust. OUH comprises four teaching hospitals in Oxfordshire, United Kingdom, with approximately 1,100 beds in total, serving a population of ~755,000 and providing specialist services to the surrounding area.

We extracted data on all adult inpatients (≥16 years) between 01 November 2021 and 31 October 2024, who were admitted under NHS patient classification codes for ordinary admissions. Day cases, regular day or night admissions were excluded as their expected LoS was predetermined. Patients admitted for obstetric or paediatric care were excluded due to the use of a different EHR system and/or discharge pathway for those specialties. Admissions were classified as elective (scheduled in advance) or emergency (admitted via the Emergency Department or other emergency units) based on the admission codes.

Clinician predictions were made by individual clinicians or during multi-disciplinary team meetings and recorded in the EHR. Not all patients had predictions documented, as this was not prioritised in all areas of the hospitals.

#### Features

To build prediction models, we used the same feature set as previously described^24^.

A total of 1,152 features were created and grouped into 16 feature categories:

1. **Index date (4 features):**

- Number of days since 1 November 2021
- Index date weekday
- Index date month
- Number of hours since admission

2. **Demographics (13 features):**

- Age
- Sex
- BMI, height, weight
- Ethnic group (white, mixed, Asian or Asian British, Black or Black British, other ethnic group, not stated/not known ethnic group)
- IMD score
- Prior mean, max, min, median, sd length of stay by postcode district

3. **Comorbidities (52 features):**

- Charlson comorbidity index, raw and age-adjusted
- Elixhauser comorbidity score, raw and age-adjusted
- Individual comorbidities in Charlson score (acute myocardial infarction, cerebral vascular disease, congestive heart failure, connective tissue disorder, dementia, diabetes, liver disease, peptic ulcer, peripheral vascular disease, pulmonary disease, cancer, diabetes complications, paraplegia, renal disease, metastatic cancer, severe liver disease, HIV)
- Individual comorbidities in Elixhauser score (congestive heart failure, cardiac arrhythmias, valvular disease, pulmonary circulation disorders, peripheral vascular disorders, hypertension uncomplicated, paralysis, other neurological disorders chronic pulmonary disease, diabetes uncomplicated, diabetes complicated, hypothyroidism, renal failure, liver disease, peptic ulcer disease, HIV, lymphoma, metastatic cancer, solid tumour without metastasis, rheumatoid arthritis, coagulopathy, obesity, weight loss, fluid and electrolyte disorders, blood loss anaemia, deficiency anaemia, alcohol abuse, drug abuse, psychoses, depression, hypertension complicated)

All comorbidities are based on diagnostic codes in previous year before the current admission.

4. **Current admission (7 features):**

- Admission time/source features:
- daytime (0 to 24 hours)
- admission weekday (Monday to Sunday)
- admission month (January to December)
- admission source (usual place of residence, non-NHS institutional care, other NHS Provider)
- Admission specialty:

Acute, emergency and geriatric medicine; Acute and general surgery; Trauma and orthopaedics; Critical care; Medical subspecialty; Surgical subspecialty; Others.

- Specialty at index date
- Number of each specialty in current admission
- Number of unique specialties admitted within 365 days before the index date
- Number of new specialties under within last 24/48 hours

5. **Ward stay (4 features):**

- Number of new wards within 24/48 hours
- The current ward is ICU
- Hours elapsed since the current ward starts

6. **Diagnosis (8 features):**

Length of stay statistics for previous admissions for all patients, by Standardised Hospital Mortality Index (SHMI) category

- LOS characteristics of SHMI diagnosis categories: Historic mean/median/maximum/minimum/SD of the LOS of patients within the same SHMI diagnostic category

7. **Discharge planning (3 features):**

- Physiotherapy referral within 24/48 hours before index date/ within 365 days before current admission date

8. **Procedures (21 features):**

- Had procedure within 24/48 hours before index date/within current admission
- Number of procedures 24/48 hours before index date/within current admission*
- Time elapsed since most recent procedure before the index date, days
- Had procedure within 365 days before current admission date
- Number of procedures within 365 days before the current admission date*

* Procedures were identified using OPCS (Operating Procedure Codes Supplement) codes. We excluded modifying codes starting with ‘Y’ and ‘Z’ to make sure each procedure was counted just once.

9. **Antibiotics prescriptions (73 features):**

- Current antibiotic use within 24/48 hours before the index date
- New antibiotics within 24/48 hours before the index date
- Antibiotics completed within 24/48 hours before index date
- Any antibiotics used within the current admission
- Duration of antibiotics used within current admission
- Count of unique antibiotics in the current admission
- Currently on a specific antibiotics agent (~60 antibiotics)

10. **Medication (36 features):**

- Use of intravenous fluids/intravenous medication/oral medication/nebulised medication/inhalation medication within 24/48 hours before index date/within current admission
- Volume of intravenous fluids within 24/48 hours before index date/within current admission
- Count of intravenous medication/oral medication/nebulised medication/inhalation medication within 24/48 hours before index date/within current admission
- Use of intravenous/oral strong opiates within 24/48 hours before index date/within current admission

11. **Microbiology tests (6 features):**

- Extended Spectrum Beta-Lactamase (ESBL)/ Carbapenemase-producing. Enterobacteriaceae (CPE) isolated in the current admission
- Vancomycin-resistant enterococci (VRE) isolated in the current admission
- Methicillin-resistant Staphylococcus aureus (MRSA) isolated in the current admission
- ESBL/CPE isolated in the last 365 days
- VRE isolated in last 365 days
- MRSA isolated in last 365 days
- Positive blood culture results within 24/48 hours before index date/within current admission

12. **Radiology investigation (9 features):**

- Had radiology-based procedure within 24/48 hours before index date/within current admission
- Number of radiology-based procedures within 24/48 hours before index date/within current admission
- Had radiology-based procedure within 365 days before current admission date
- Number of radiology-based procedures within 365 days before the current admission date
- Time elapsed since most recent radiology procedure within 365 days before the index date

13. **Readmissions and previous hospital stay (22 features):**

- Readmissions
- Current admission is early readmission: ≤30 days from a previous hospitalization event
- Current admission is late readmission: >30 to ≤180 days from a previous hospitalization event
- Number of early 30-day readmissions within 365 days before the index date
- Time elapsed from most recent early 30-day readmission within 365 days before the index date, days
- Previous length of stay
- Number of admissions within 30/90/365 days before the index date
- Cumulative LOS within 30/90/365 days before the index date
- Mean/Maximum/Minimum/SD LOS per admission within 30/90/365 days before the current admission date

14. **Hospital capacity factors (23 features):**

- The median/mean/maximum/minimum/SD LOS of the current ward
- The current number of inpatients in the hospital
- Current inpatients with LOS to date of >= 7 days, 14 days, 28 days
- Proportion of current inpatients with LOS to date of >= 7 days, 14 days, 28 days
- Number of admissions within the last 24hr, 48hr, 7d, 28d
- Number of discharges within the last 24h, 48h, 7d, 28d
- The mean LOS for all discharges in the 7d, 14d, 28d before the index date

15. **Vital signs (135 features):**

- Mean / Max / Min / SD of each vital sign measurements (see below) within 24/48 hours before index date/within current admission
- Number of vital signs measurements within 24/48 hours before index date/within current admission

**List of vital signs:** heart rate, respiratory rate, systolic blood pressure, diastolic blood pressure, temperature, oxygen saturation, O2 L/min, O2 delivery device, AVPU score, NEWS2 score, NEWS2 score alternative (missing oxygen device = Room air)

16. **Laboratory tests (736 features):**

- Mean / Max / Min / SD of each laboratory test measurements within 24/48 hours before index date/within current admission
- Number of laboratory test measurements within 24/48 hours before index date/within current admission/within 365 days before the index date

**List of laboratory tests:**

- **Complete blood counts:** Haemoglobin, Haematocrit, Mean Cell Volume, White Cell Count, Platelets, Neutrophils, Lymphocytes, Eosinophils, Monocytes, Basophils
- **Renal functions:** Creatinine, Urea, Potassium, Sodium, EGFR, Bicarbonate
- **Inflammatory:** C-reactive protein, Erythrocyte Sedimentation Rate, Creatinine kinase
- **Liver functions:** Alkaline phosphatase, Aspartate aminotransferase, Alanine transaminase, Albumin, Bilirubin, Amylase, Gamma-glutamyl Transferase
- **Bone/electrolytes profiles:** Adjusted calcium, Magnesium, Phosphate
- **Clotting:** Activated partial thromboplastin time, Prothrombin time, D_dimer
- **Endocrine:** Thyroid-stimulating hormone, HbA1c, Glucose, Prostate-specific antigen
- **Haematinics:** Ferritin, Iron, Transferrin, B12, Folate
- **Others:** Lactate dehydrogenase, Troponin, Total Ig
- **Blood gases:** Base excess, Partial pressure of oxygen, Partial pressure of carbon dioxide, Lactate, Arterial blood pH
- **Lipids:** Triglycerides, High-density lipoprotein cholesterol, Total cholesterol, Low-density lipoprotein cholesterol

#### Preprocessing

Implausible extreme outliers (e.g. height 10m, temperature 20 ^o^C) were treated as missing values as they usually represented errors in data entry. The proportion of missingness for features ranged from 0 to 88% for individual features. Categorical features were imputed using the mode, while numerical features were imputed using the median. Categorical features were one-hot encoded, and numerical features were normalised using Min-Max scaling. The preprocessing pipeline was fitted on the training dataset and applied to the test dataset.

#### Prediction outcome

We predicted in-hospital mortality and future hospital LoS at an index date and time for all patients currently in the hospital. The in-hospital mortality was extracted as a binary event and determined by discharge method code and discharge destination code, with the discharge date used as the mortality date. Future hospital LoS were categorised into quartiles, reflecting clinically relevant durations of future stay:

- Elective admissions: Q1: 0-1, Q2: 2-3, Q3: 4-10, Q4: >10 days.
- Emergency admissions: Q1: 0-2, Q2: 3-6, Q3: 7-14, Q4: >14 days.

Predictions were made daily at 12 pm to align with the end of typical hospital ward rounds. Each patient contributed once to the dataset each day during their hospital admission. Separate models were built for emergency and elective admissions as predictive features plausibly differ by the admission type. Models were trained using data from the first two years (01 November 2021 to 31 October 2023) and evaluated using data from the final year (01 November 2023 to 31 October 2024).

#### Single-task models

In our single-task model analysis, we used binary extreme gradient boosting (XGB) to predict mortality, and multiclass XGB to predict hospital LoS. We randomly split the training data, using 80% for main model training. Within this, we used Bayesian optimization with Tree-based Parzen estimators (TPE) for hyperparameter tuning, maximizing the area under the receiver operating curve (AUROC) through 5-fold cross-validation. Hyperparameters tuned included learning rate, the number of trees, the fraction of features to use, the fraction of observations to subsample at each step, the maximum number of nodes allowed from the root to the farthest leaf of a tree, the minimum weight required to create a new node in a tree, the minimum loss reduction required to make a split, L1 regularisation term on weight, and L2 regularisation term on weight. Class imbalance was addressed using scale_pos_weight for mortality and sample_weight for LoS.

We used the remaining 20% of the training dataset as validation data for feature selection, calibrating the predicted probability from XGB models using isotonic regression, and determining the best threshold for predicting mortality by optimising the F1 score (jointly maximising precision and recall). XGB models were initially trained with all 1,152 features. Feature importance was determined using the SHapley Additive exPlanations (SHAP) values^25^ for each feature in the training dataset. Then, the top-ranked 200 features were retained from each full model as the optimal number of features, as in a previous study we found that model performance increased as more features were included at the expense of increased training time, but largely plateaued with ≥200 features^24^.

We then compared the XGB models to logistic regression (LR) and multilayer perceptron (MLP) models, using the same 200 features selected for the XGB models. We built binary LR for mortality, and multinomial LR for LoS as baseline comparators, with the default L2 regularisation (C=1) and a balanced class weight to account for class imbalance. The MLP model used fully connected layers with dropout, binary cross entropy loss with sigmoid function (BCEwithlogitsloss) for mortality and cross-entropy loss function for LoS. We used the Adam optimizer and early stopping with a patience of 10 epochs to avoid overfitting. The number of layers and dropout ratio were manually tuned. Learning rate and weight decay were tuned using Grid search. We applied weights in the loss function to deal with class imbalance.

#### Multiclass classification model

We then fitted an XGB model similarly to above by combining mortality and LoS quartiles into a single-task multiclass model with seven outcome categories:

- Elective: discharged alive within 0-1 days (Q1), 2-3 days (Q2), or 4-10 days (Q3); died in hospital within 0-1 days, 2-3 days, or 4-10 days; or remained in hospital >10 days (Q4).
- Emergency: discharged alive within 0-2 days (Q1), 3-6 days (Q2), or 7-14 days (Q3); died in hospital within 0-2 days, 3-6 days, or 7-14 days; or remained in hospital >14 days (Q4).

#### Multi-task learning model

MTL uses shared features to enhance performance and efficiency across tasks^18^. We used an MLP architecture based on the same feature set as the single-task models to predict mortality and LoS simultaneously. The same loss functions and hyperparameters were used as in the single-task models. To properly combine the task-specific losses, we compared different loss weighting strategies, including uniform weighting (with different weights), random loss weighting^26^, and uncertainty weighting^27^.

#### Performance assessment

For binary tasks, we evaluated the model performance using accuracy, sensitivity (recall), specificity, positive predictive value (PPV, precision), negative predictive value (NPV), F1 score (harmonic mean of precision and recall), area under the receiver operating curve (AUROC), and area under the precision-recall curve (AUPRC). For multiclass tasks, we evaluated accuracy, macro-averaged precision, recall, F1 score, AUROC, and AUPRC. Macro-averaging ensured equal weighting across classes (i.e. an unweighted mean performance)^28^. 95% confidence intervals (CIs) were estimated using bootstrap.

We compared the performance of the LoS models with clinician predictions made within the testing period. As clinician predictions were made by clinicians at random timepoints throughout the day, for comparisons a separate model prediction was made, using the time of the prediction, rather than 12 pm, as the index date and time for data extract.

#### Software

Data processing and analyses were performed in Python 3.11 using the following packages: numpy (version 1.26.4), pandas (version 2.2.0), scipy (version 1.12.0), scikit-learn (version 1.4.1), xgboost (version 2.0.3), optuna (version 3.6.1), torch (version 2.3.0) and in R (version 4.3.2) using the following packages: cowplot (version 1.1.1), timeDate (version 4021.104), ggplot2 (version 3.4.4), and tidyverse (version 1.3.2).

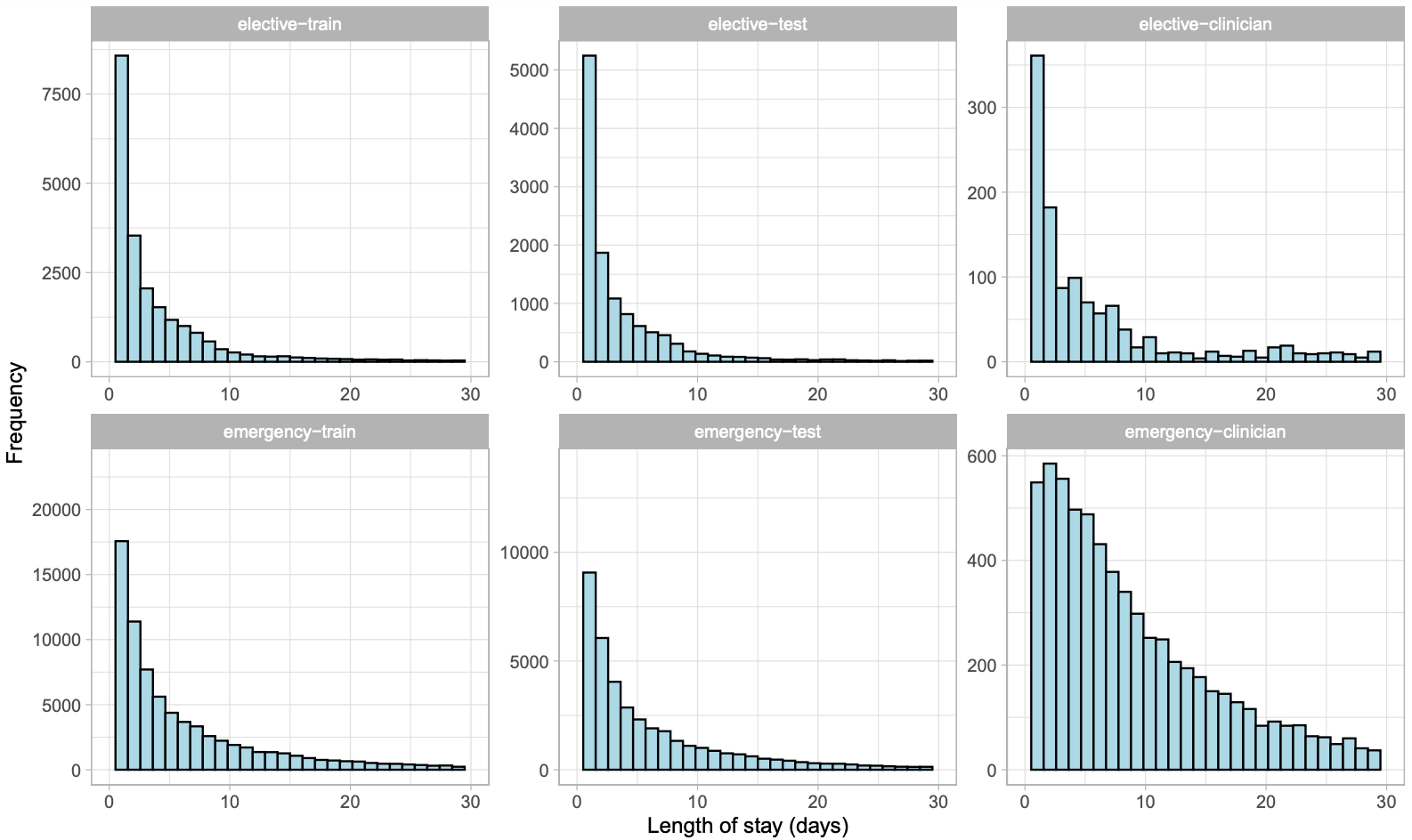

**Figure S1. Distribution of length of stay (from admission to discharge) for elective and emergency patients.** Distribution is similar between training and test data. Admissions with clinician predictions have a longer length of stay. Length of stay is censored at 30 days for better visualisation (421, 169, and 63 elective admissions, and 3561, 1462, and 539 emergency admissions in training, test, and clinician data had a length of stay >30 days).

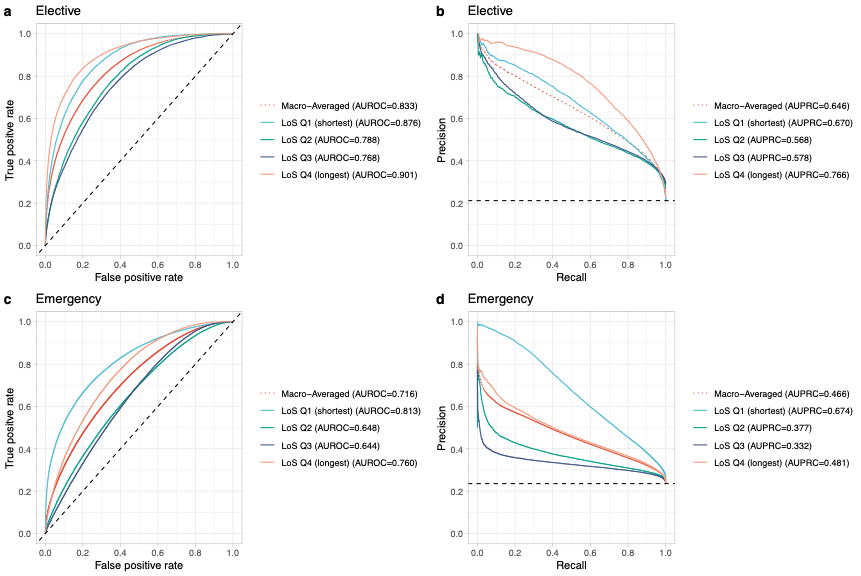

**Figure S2.** **Model performance of the extreme gradient boosting models predicting future hospital length of stay in the test dataset.** a) Area under the receiver operating curve (AUROC) for elective admissions. b) Area under the precision-recall curve (AUPRC) for elective admissions. c) AUC for emergency admissions. d) AUPRC for emergency admissions. Q1: below 25% quantile (0-1 days for elective, 0-2 days for emergency); Q2: 25% to 50% quantile (2-3 days for elective, 3-6 days for emergency); Q3: 50% to 75% quantile (4-10 days for elective, 7-14 days for emergency); Q4: above 75% quantile (>10 days for elective, >14 days for emergency).

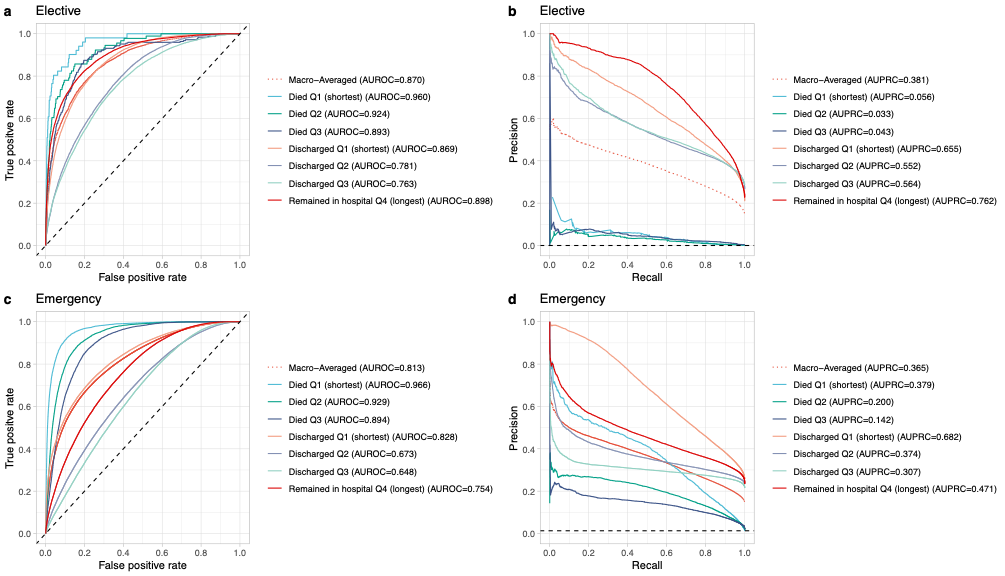

**Figure S3. Model performance of the extreme gradient boosting models predicting combined in-hospital mortality and future hospital length of stay in the test dataset.** a) Area under the receiver operating curve (AUROC) for elective admissions. b) Area under the precision-recall curve (AUPRC) for elective admissions. c) AUC for emergency admissions. d) AUPRC for emergency admissions. Class 0: discharged alive within 0-1 day for elective, and 0-2 days for emergency; Class 1: discharged alive within 2-3 days for elective, and 3-6 days for emergency; Class 2: discharged alive within 4-10 days for elective, and 7-14 days for emergency; Class 3: died in hospital within 0-1 day for elective, and 0-2 days for emergency; Class 4: died in hospital within 2-3 days for elective, and 3-6 days for emergency; Class 5: died in hospital within 4-10 days for elective, and 7-14 days for emergency; Class 6: remained in hospital >10 days for elective, and >14 days for emergency.

| Admission type | Model | Accuracy | Sensitivity/Recall | Specificity | PPV/Precision | NPV | F1-score | AUROC | AUPRC |
| --- | --- | --- | --- | --- | --- | --- | --- | --- | --- |
| Mortality |  |  |  |  |  |  |  |  |  |
| Elective | XGB | 0.980 (0.979-0.981) | 0.225 (0.197-0.25) | 0.991 (0.990-0.991) | 0.262 (0.232-0.288) | 0.989 (0.988-0.989) | 0.242 (0.215-0.264) | 0.917 (0.910-0.924) | 0.219 (0.146-0.268) |
|  | LR | 0.906 (0.904-0.908) | 0.585 (0.554-0.616) | 0.911 (0.909-0.913) | 0.088 (0.08-0.094) | 0.993 (0.993-0.994) | 0.152 (0.140-0.162) | 0.876 (0.865-0.888) | 0.204 (0.187-0.223) |
|  | MLP | 0.926 (0.924-0.928) | 0.662 (0.632-0.686) | 0.930 (0.928-0.932) | 0.122 (0.114-0.129) | 0.995 (0.994-0.995) | 0.206 (0.193-0.217) | 0.922 (0.916-0.929) | 0.206 (0.181-0.227) |
|  | Multi-task | 0.924 (0.921-0.925) | 0.649 (0.616-0.677) | 0.928 (0.925-0.929) | 0.116 (0.107-0.122) | 0.995 (0.994-0.995) | 0.196 (0.182-0.207) | 0.915 (0.908-0.921) | 0.191 (0.167-0.215) |
| Emergency | XGB | 0.927 (0.926-0.927) | 0.571 (0.565-0.577) | 0.957 (0.956-0.958) | 0.532 (0.526-0.536) | 0.963 (0.962-0.964) | 0.551 (0.546-0.556) | 0.924 (0.922-0.926) | 0.569 (0.562-0.575) |
|  | LR | 0.916 (0.916-0.917) | 0.556 (0.550-0.562) | 0.947 (0.947-0.948) | 0.475 (0.469-0.479) | 0.961 (0.961-0.962) | 0.512 (0.507-0.517) | 0.907 (0.905-0.909) | 0.494 (0.487-0.501) |
|  | MLP | 0.919 (0.918-0.92) | 0.563 (0.557-0.57) | 0.949 (0.948-0.95) | 0.488 (0.482-0.493) | 0.962 (0.961-0.963) | 0.523 (0.517-0.528) | 0.911 (0.909-0.913) | 0.523 (0.517-0.531) |
|  | Multi-task | 0.922 (0.921-0.923) | 0.561 (0.555-0.567) | 0.953 (0.952-0.954) | 0.506 (0.501-0.511) | 0.962 (0.961-0.963) | 0.532 (0.527-0.537) | 0.914 (0.912-0.915) | 0.542 (0.535-0.549) |
| LoS |  |  |  |  |  |  |  |  |  |
| Elective | XGB | 0.579 (0.575-0.583) | 0.579 (0.576-0.584) |  | 0.605 (0.602-0.609) |  | 0.587 (0.584-0.591) | 0.833 (0.831-0.835) | 0.645 (0.642-0.65) |
|  | LR | 0.527 (0.523-0.53) | 0.533 (0.529-0.536) |  | 0.535 (0.532-0.539) |  | 0.534 (0.531-0.537) | 0.786 (0.784-0.788) | 0.557 (0.552-0.56) |
|  | MLP | 0.553 (0.549-0.556) | 0.567 (0.563-0.57) |  | 0.553 (0.549-0.557) |  | 0.557 (0.552-0.56) | 0.811 (0.809-0.813) | 0.602 (0.597-0.606) |
|  | Multi-task | 0.545 (0.541-0.549) | 0.561 (0.557-0.564) |  | 0.549 (0.545-0.553) |  | 0.549 (0.545-0.552) | 0.807 (0.805-0.809) | 0.594 (0.589-0.598) |
| Emergency | XGB | 0.446 (0.444-0.448) | 0.441 (0.44-0.443) |  | 0.431 (0.43-0.433) |  | 0.434 (0.432-0.436) | 0.716 (0.715-0.718) | 0.466 (0.465-0.468) |
|  | LR | 0.416 (0.414-0.418) | 0.412 (0.41-0.413) |  | 0.397 (0.396-0.399) |  | 0.395 (0.393-0.396) | 0.683 (0.682-0.684) | 0.421 (0.419-0.422) |
|  | MLP | 0.434 (0.432-0.435) | 0.431 (0.43-0.433) |  | 0.424 (0.422-0.425) |  | 0.425 (0.423-0.426) | 0.702 (0.701-0.703) | 0.446 (0.444-0.447) |
|  | Multi-task | 0.433 (0.431-0.435) | 0.428 (0.426-0.43) |  | 0.422 (0.420-0.424) |  | 0.424 (0.422-0.425) | 0.703 (0.702-0.704) | 0.446 (0.445-0.448) |
| Combined mortality+LoS | |  |  |  |  |  |  |  |  |
| Elective | XGB | 0.572 (0.568-0.575) | 0.344 (0.334-0.357) |  | 0.358 (0.342-0.376) |  | 0.349 (0.337-0.361) | 0.870 (0.865-0.876) | 0.381 (0.376-0.392) |
|  | LR | 0.517 (0.513-0.52) | 0.31 (0.303-0.317) |  | 0.373 (0.311-0.451) |  | 0.315 (0.303-0.329) | 0.838 (0.831-0.845) | 0.348 (0.336-0.366) |
| Emergency | XGB | 0.435 (0.433-0.436) | 0.331 (0.329-0.334) |  | 0.366 (0.360-0.372) |  | 0.328 (0.326-0.331) | 0.813 (0.812-0.814) | 0.365 (0.363-0.368) |
|  | LR | 0.410 (0.409-0.412) | 0.303 (0.300-0.306) |  | 0.351 (0.346-0.356) |  | 0.3 (0.297-0.303) | 0.789 (0.788-0.790) | 0.330 (0.327-0.332) |

**Table S1.** **Model performance of single-task and multi-task models predicting in-hospital mortality and future hospital length of stay (LoS).** Separate models for elective and emergency admissions were trained using the top 200 most predictive features selected from the XGB models. Mean with 95% confidence intervals are calculated using bootstrap. XGB: extreme gradient boosting; LR: logistic regression; MLP: multi-layer perceptron; PPV: positive predictive value; NPV: negative predictive value; AUROC: area under the receiver operating curve; AUPRC: area under the precision-recall curve.

| Admission type | Data | Accuracy | Sensitivity/Recall | Specificity | PPV/Precision | NPV | F1-score | AUROC | AUPRC |
| --- | --- | --- | --- | --- | --- | --- | --- | --- | --- |
| Mortality |  |  |  |  |  |  |  |  |  |
| Elective | train | 0.999 (0.999-0.999) | 0.946 (0.934-0.957) | 1.0 (1.0-1.0) | 1.0 (1.0-1.0) | 0.999 (0.999-0.999) | 0.972 (0.966-0.978) | 0.999 (0.998-0.999) | 0.988 (0.983-0.992) |
| Elective | validation | 0.971 (0.969-0.973) | 0.269 (0.231-0.309) | 0.987 (0.986-0.988) | 0.320 (0.278-0.367) | 0.983 (0.982-0.985) | 0.292 (0.257-0.334) | 0.944 (0.938-0.949) | 0.296 (0.226-0.357) |
| Elective | test | 0.980 (0.979-0.981) | 0.225 (0.197-0.250) | 0.991 (0.99-0.991) | 0.262 (0.232-0.288) | 0.989 (0.988-0.989) | 0.242 (0.215-0.264) | 0.917 (0.910-0.924) | 0.219 (0.146-0.268) |
| Emergency | train | 0.979 (0.978-0.979) | 0.878 (0.875-0.88) | 0.988 (0.988-0.988) | 0.870 (0.867-0.873) | 0.989 (0.988-0.989) | 0.874 (0.872-0.876) | 0.990 (0.990-0.990) | 0.943 (0.942-0.945) |
| Emergency | validation | 0.916 (0.914-0.917) | 0.559 (0.550-0.566) | 0.949 (0.948-0.95) | 0.503 (0.493-0.511) | 0.959 (0.958-0.960) | 0.529 (0.521-0.536) | 0.911 (0.908-0.914) | 0.549 (0.539-0.558) |
| Emergency | test | 0.927 (0.926-0.927) | 0.571 (0.565-0.577) | 0.957 (0.956-0.958) | 0.532 (0.526-0.536) | 0.963 (0.962-0.964) | 0.551 (0.546-0.556) | 0.924 (0.922-0.926) | 0.569 (0.562-0.575) |
| LoS |  |  |  |  |  |  |  |  |  |
| Elective | train | 0.838 (0.836-0.841) | 0.833 (0.831-0.836) |  | 0.838 (0.836-0.841) |  | 0.835 (0.833-0.838) | 0.963 (0.962-0.963) | 0.897 (0.895-0.899) |
| Elective | validation | 0.573 (0.568-0.580) | 0.576 (0.570-0.583) |  | 0.601 (0.595-0.608) |  | 0.579 (0.573-0.585) | 0.836 (0.833-0.839) | 0.646 (0.639-0.653) |
| Elective | test | 0.579 (0.575-0.583) | 0.579 (0.576-0.584) |  | 0.605 (0.602-0.609) |  | 0.587 (0.584-0.591) | 0.833 (0.831-0.835) | 0.645 (0.642-0.650) |
| Emergency | train | 0.591 (0.59-0.592) | 0.586 (0.584-0.587) |  | 0.582 (0.581-0.583) |  | 0.582 (0.580-0.583) | 0.831 (0.831-0.832) | 0.651 (0.650-0.652) |
| Emergency | validation | 0.447 (0.445-0.449) | 0.444 (0.441-0.446) |  | 0.437 (0.435-0.439) |  | 0.439 (0.437-0.442) | 0.718 (0.717-0.720) | 0.470 (0.468-0.473) |
| Emergency | test | 0.446 (0.444-0.448) | 0.441 (0.440-0.443) |  | 0.431 (0.430-0.433) |  | 0.434 (0.432-0.436) | 0.716 (0.715-0.718) | 0.466 (0.465-0.468) |

**Table S2.** **Model performance of the extreme gradient boosting (XGB) model predicting in-hospital mortality and future hospital length of stay (LoS) in the training, validation, and test dataset.** Mean with 95% confidence intervals are calculated using bootstrap. PPV: positive predictive value; NPV: negative predictive value; AUROC: area under the receiver operating curve; AUPRC: area under the precision-recall curve.

| LoS quantile | Length of stay | Number of patient-days | Outcome | Number of patient days |
| --- | --- | --- | --- | --- |
| Elective |  |  |  |  |
| Q1 | 0-1 days | 37,472 (20.1%) | Died | 158 (0.1%) |
|  |  |  | Discharged | 37,314 (20.0%) |
| Q2 | 2-3 days | 48,324 (25.9%) | Died | 280 (0.2%) |
|  |  |  | Discharged | 48,044 (25.7%) |
| Q3 | 4-10 days | 54,096 (29.0%) | Died | 733 (0.4%) |
|  |  |  | Discharged | 53,363 (28.6%) |
| Q4 | >10 days | 46,735 (25.0%) | Died+Discharged | 46,735 (25.0%) |
| Emergency |  |  |  |  |
| Q1 | 0-2 days | 247,517 (25.8%) | Died | 12,449 (1.3%) |
|  |  |  | Discharged | 235,068 (24.5%) |
| Q2 | 3-6 days | 237,967 (24.8%) | Died | 18,489 (1.9%) |
|  |  |  | Discharged | 219,478 (22.9%) |
| Q3 | 7-14 days | 228,486 (23.8%) | Died | 22,104 (2.3%) |
|  |  |  | Discharged | 206,382 (21.5%) |
| Q4 | >14 days | 246,384 (25.6%) | Died+Discharged | 246,384 (25.6%) |

**Table S3. Number of patient days in each class by admission type and outcome.** Length of stay (LoS) quartiles (Q1-Q4) were used in the single-task and multi-task models for predicting future hospital length of stay. Each quartile was further divided into died in hospital and discharged alive to be used in the multi-class classification model for predicting combined in-hospital death and hospital length of stay.

| Admission type | Weight | Accuracy | Sensitivity/Recall | Specificity | PPV/Precision | NPV | F1-score | AUROC | AUPRC |
| --- | --- | --- | --- | --- | --- | --- | --- | --- | --- |
| Elective | Mortality: LoS |  |  |  |  |  |  |  |  |
| Mortality | 10:1 | 0.929 (0.927-0.931) | 0.643 (0.609-0.668) | 0.933 (0.931-0.935) | 0.123 (0.115-0.13) | 0.994 (0.994-0.995) | 0.207 (0.194-0.217) | 0.919 (0.913-0.926) | 0.201 (0.175-0.223) |
| Mortality | 1:1 | 0.958 (0.957-0.96) | 0.464 (0.432-0.489) | 0.966 (0.964-0.967) | 0.164 (0.151-0.175) | 0.992 (0.991-0.993) | 0.243 (0.226-0.258) | 0.914 (0.908-0.921) | 0.180 (0.157-0.199) |
| Mortality | 1:10 | 0.924 (0.921-0.925) | 0.649 (0.616-0.677) | 0.928 (0.925-0.929) | 0.116 (0.107-0.122) | 0.995 (0.994-0.995) | 0.196 (0.182-0.207) | 0.915 (0.908-0.921) | 0.191 (0.167-0.215) |
| Mortality | uncertainty | 0.938 (0.936-0.94) | 0.596 (0.565-0.623) | 0.943 (0.941-0.945) | 0.132 (0.124-0.141) | 0.994 (0.993-0.994) | 0.216 (0.204-0.228) | 0.919 (0.913-0.927) | 0.213 (0.186-0.233) |
| Mortality | random | 0.973 (0.972-0.975) | 0.322 (0.295-0.346) | 0.983 (0.982-0.984) | 0.215 (0.194-0.231) | 0.99 (0.989-0.991) | 0.258 (0.236-0.275) | 0.915 (0.909-0.921) | 0.178 (0.156-0.197) |
| LoS | 10:1 | 0.392 (0.389-0.396) | 0.403 (0.399-0.407) |  | 0.392 (0.388-0.396) |  | 0.395 (0.391-0.399) | 0.659 (0.657-0.662) | 0.401 (0.398-0.405) |
| LoS | 1:1 | 0.460 (0.456-0.464) | 0.485 (0.482-0.488) |  | 0.466 (0.462-0.47) |  | 0.456 (0.452-0.46) | 0.748 (0.745-0.75) | 0.498 (0.494-0.502) |
| LoS | 1:10 | 0.545 (0.541-0.549) | 0.561 (0.557-0.564) |  | 0.549 (0.545-0.553) |  | 0.549 (0.545-0.552) | 0.807 (0.805-0.809) | 0.594 (0.589-0.598) |
| LoS | uncertainty | 0.460 (0.456-0.463) | 0.478 (0.475-0.481) |  | 0.468 (0.465-0.472) |  | 0.462 (0.459-0.465) | 0.735 (0.733-0.737) | 0.480 (0.476-0.483) |
| LoS | random | 0.464 (0.460-0.468) | 0.487 (0.483-0.491) |  | 0.465 (0.461-0.469) |  | 0.462 (0.458-0.466) | 0.746 (0.743-0.749) | 0.496 (0.492-0.500) |
| Emergency | Mortality: LoS |  |  |  |  |  |  |  |  |
| Mortality | 10:1 | 0.920 (0.919-0.92) | 0.582 (0.576-0.589) | 0.948 (0.948-0.949) | 0.491 (0.485-0.497) | 0.964 (0.963-0.964) | 0.533 (0.527-0.538) | 0.913 (0.911-0.915) | 0.538 (0.531-0.545) |
| Mortality | 1:1 | 0.923 (0.922-0.924) | 0.560 (0.553-0.567) | 0.954 (0.953-0.955) | 0.510 (0.504-0.515) | 0.962 (0.961-0.963) | 0.533 (0.527-0.539) | 0.914 (0.912-0.916) | 0.54 (0.533-0.546) |
| Mortality | 1:10 | 0.922 (0.921-0.923) | 0.561 (0.555-0.567) | 0.953 (0.952-0.954) | 0.506 (0.501-0.511) | 0.962 (0.961-0.963) | 0.532 (0.527-0.537) | 0.914 (0.912-0.915) | 0.542 (0.535-0.549) |
| Mortality | uncertainty | 0.921 (0.92-0.922) | 0.576 (0.569-0.582) | 0.950 (0.950-0.951) | 0.499 (0.492-0.504) | 0.963 (0.963-0.964) | 0.534 (0.529-0.539) | 0.914 (0.912-0.916) | 0.543 (0.536-0.550) |
| Mortality | random | 0.919 (0.918-0.92) | 0.582 (0.576-0.588) | 0.948 (0.947-0.948) | 0.487 (0.481-0.493) | 0.964 (0.963-0.964) | 0.530 (0.524-0.536) | 0.914 (0.913-0.916) | 0.539 (0.532-0.546) |
| LoS | 10:1 | 0.369 (0.367-0.37) | 0.365 (0.364-0.367) |  | 0.343 (0.341-0.345) |  | 0.341 (0.339-0.343) | 0.64 (0.638-0.641) | 0.376 (0.374-0.377) |
| LoS | 1:1 | 0.426 (0.425-0.428) | 0.423 (0.421-0.425) |  | 0.411 (0.409-0.413) |  | 0.413 (0.411-0.415) | 0.694 (0.693-0.695) | 0.433 (0.432-0.435) |
| LoS | 1:10 | 0.433 (0.431-0.435) | 0.428 (0.426-0.430) |  | 0.422 (0.420-0.424) |  | 0.424 (0.422-0.425) | 0.703 (0.702-0.704) | 0.446 (0.445-0.448) |
| LoS | uncertainty | 0.429 (0.427-0.431) | 0.426 (0.424-0.428) |  | 0.414 (0.412-0.417) |  | 0.416 (0.414-0.418) | 0.695 (0.694-0.696) | 0.435 (0.433-0.437) |
| LoS | random | 0.411 (0.41-0.413) | 0.408 (0.406-0.409) |  | 0.388 (0.386-0.389) |  | 0.387 (0.385-0.389) | 0.679 (0.678-0.680) | 0.416 (0.414-0.417) |

**Table S4.** **Model performance of the multi-task model predicting in-hospital mortality and future hospital length of stay (LoS) in the test dataset.** Different weighting strategies are compared, including uniform weighting (weights for mortality: LoS, 10:1, 1:1, 1:10), uncertainty weighting, and random weighting. Shaded rows are the final model with the best performance. Mean with 95% confidence intervals are calculated using bootstrap. PPV: positive predictive value; NPV: negative predictive value; AUROC: area under the receiver operating curve; AUPRC: area under the precision-recall curve.
